# Unequal Lives: A Sociodemographic Analysis of Covid19 Transmission and Mortality in India

**DOI:** 10.1101/2020.09.06.20189506

**Authors:** Soham Dibyachintan, Priyanka Nandy, Kalyan Das, Sai Vinjanampathy, Mithun K. Mitra

**Author notes:** Email addresses:* (Sai Vinjanampathy), (Mithun K. Mitra).

## Abstract

The hierarchy of social structures shape, in very particular and measurable ways, the differential impact that a disease has on different parts of society. In this study, we use district-level disease data to perform an ecological analysis of Covid19 outcomes in India vis a vis the local socioeconomic gradient. Average doubling times and case fatality ratios have been quantified as measures of transmission and mortality, respectively, and association analysis performed with twenty variables of socioeconomic vulnerability. Persistent patterns are observed between disease outcome and social inequality, linking poor living conditions to a faster spread, an elderly populace to a slower spread, and both a college education and the presence of medical facilities to low fatality rates.

## 1. Introduction

In the spread of an infectious disease, the human population is as much the victim as it is the vector. But not all vectors are created equal. When mapped on a bounded geographical space, particularly a diverse one, the path of an infectious disease manifests as a barium test for the anatomy of its inequalities^1,2^. Indeed, while the WHO Commission on the Social Determinants of Health had been founded in 2005, the pathogenic behaviour of social inequalities has been seminally established in literature for nearly two centuries^3,4^. Contemporary studies have demonstrated conclusively that the gradient of localised inequalities - indexed by social marginality^5–8^, employment status^9–11^, educational attainment^12^, wealth/asset gap^13,14^, access to social^15,16^ or public support systems^17,18^, and mortality patterns^10,19–21^– affect the differential impact of both non-communicable^10,19,22^ and infectious^23^ diseases, and can only be addressed by evidence-based, ethical policies focused on public good^24,25^

If one goal of disease modelling is to aid the efficacy of public health responses and to eventually contribute towards more equal outcomes in health^21,26^, then it is crucial to locate the socioeconomic variables that map onto patterns of persistent transmission and mortality. In the contemporary instance of Covid19, the centrality of age-structure^27,28^ and its clinical implications^29^ in assessing spread and mortality outcomes has already been established. Age-structure, however, is only one of several demographic factors that is shaping the path of Covid19 through a community^30^. To more fully understand the impact of inequality on Covid19 outcomes - particularly in India, where even small spatial units present considerable diversity in age structure, income, housing, caste, ethnicity, religion, educational attainment, migration status, and availability of health services - a more comprehensively ecological approach is necessary. Similar analysis has yielded information as to how socioeconomic disparities may have played a role in past epidemics^31,32^. The evidence from such an analysis can help shape “smart” interventions responsive to specific lacunae at each unit, rather than a monolithic national model^33^.

While multiple modelling studies have attempted to analyse and predict the spread of Covid19 in India from the perspective of epidemiological models^34–40^, the correlation of impact and ecology has not been explored so far^1,41^. In this study, we have thus attempted to correlate the trajectory of spread and mortality of the SARS-CoV-2 virus in India, with variables of social and economic vulnerability persistent within its populace. We hope that our findings shall contribute towards a more inclusive epistemology of the virus, as well as serve as the beginnings of a repository that shall inform an evidence-based model of public health policy that is sensitive to socioeconomic gradients and adaptive to local variations of need.

## 2. Methodology and Materials

To detect local variation, we have chosen districts as our unit of spatial analysis, for it is the smallest administrative unit for which we have consistently available data. Data was obtained from the crowd-sourced database http://covid19india.org^42^. The analysis in this paper was done with data accumulated up to the 2nd of July, 2020 - the beginning of the Unlock 2 phase in India’s Covid19 containment protocol.

We used the doubling time as the measure to characterise transmission dynamics. The doubling time (τ_D_) was calculated for every district as a time series, and these values were then averaged over time to obtain the mean doubling time for every district. Note that as the recording of cases began at very different times in different districts, these time averages are necessarily over differing periods. Based on the available data, doubling times were estimated for 707 districts in India (which was the total number of districts with more than a single case at the time of the analysis). To characterise mortality, we use the Case Fatality Ratio (CFR), which was similarly calculated as a time series for all districts that recorded at least one Covid19 mortality. The CFR data was averaged over a period of 15 days preceding the last date of measurement in order to minimise fluctuations due to initial transients. Based on the available data, the CFR was estimated for 433 districts in India (there were as yet no recorded deaths in the remaining districts). Sample time series plots, for both the doubling time and the CFR are shown for two districts in Suppl. Figure S1.

For an analysis of the correlation between social vulnerability with disease transmission and mortality, we chose twenty indices from a range of socio-demographic variables from the Census of India 2011^43^. These were then organised into 6 analytical categories, as follows:

### Age structure

(i) Total population of the district; (ii) Number of children aged 0–6 years in the district; (iii) Number of elderly persons (aged 60 years or above).

### Educational attainment

(iv) Literacy rate of a district, where any person aged seven years and above who can read and write in any language is enumerated as literate; (v) Percentage of population with primary education; (vi) Percentage of population with secondary education; (vii) Percentage of population with a college education.

### Household status

(viii) Total number of households per district, including Normal, Institutional, and Houseless households; (ix) Number of houses in “good” condition, defined by the census as structurally sound and in a condition of good repair; (x) Number of houses in “dilapidated” condition, defined as either needing major structural repairs or being beyond repair; (xi) Number of households with no drainage facilities for non-latrine waste water; (xii) Number of households with no latrine available within the house; (xiii) Number of households where the source of drinking water is “away”, defined by the census as being at a distance of at least 100 metres in an urban area and 500 metres in a rural area; (xiv) Number of households with no electricity.

### Economic activity

(xv) The population of main workers - those who worked for more than 180 days in the one year preceding the date of enumeration; (xvi) The population of marginal workers - those who worked less than 180 days in the one year preceding the date of enumeration; (xvii) Non-working population - those who did not engage in any economically productive activity during the last year.

### Social marginality

(xviii) Scheduled Castes and Tribes (SC/ST) population of the district, considered the most socially marginalised and therefore economically disadvantaged; (xix) Population with disabilities, both mental and physical.

### Health Infrastructure

(xx) Availability of medical facilities.

In order to validate that the observed trends were robust, we created five categories of districts for both the doubling time (τ_D_) and CFR. Note that the number of districts at the time of Census of India 2011 was 591, and hence the analysis could only be performed for these districts. For the τ_D_ analysis, these five subsets were: (i) all districts with greater than one confirmed case (N_all_ = 578), (ii) districts with more than 50 cases (N_50_ = 488), (iii) districts with more than 70 cases (N_70_ = 452), (iv) districts with more than 100 cases (N_100_ = 403), and (v) districts with more than 150 cases (N_150_ = 323). For the CFR analysis, the five subsets were: (i) all districts with at least one confirmed death 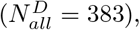, (ii) districts with at least five deaths 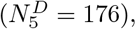, (iii) districts with at least ten deaths 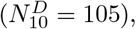, (iv) districts with at least 20 deaths 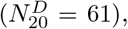, and (v) districts with at least 30 deaths 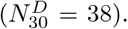. These thresholds were chosen from the distribution of cases and mortality statistics for all districts (Figure 1). A time evolution of the cumulative frequency distribution of reported cases and fatalities at landmark points in India’s national containment strategy is shown in Suppl. Figure S2.

**Figure 1:**
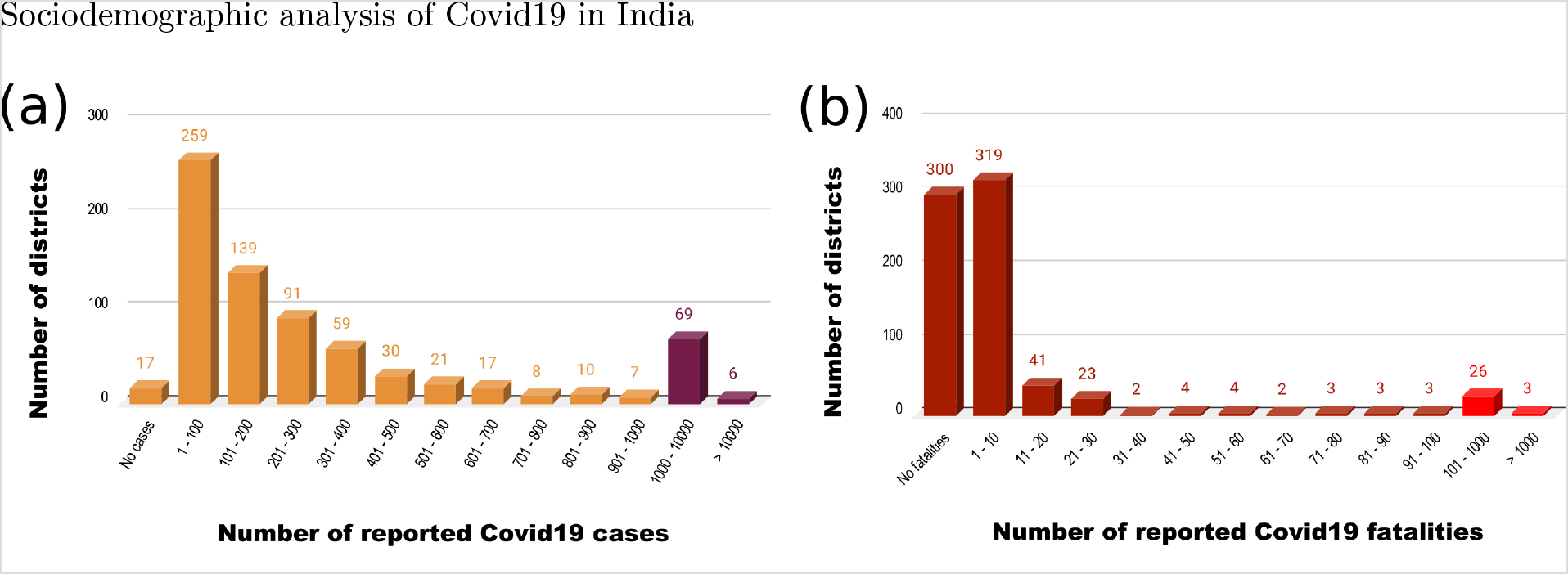
Histograms of the number of districts with the (a) number of recorded cases, and (b) number of recorded fatalities. The data is till the 2nd of July, 2020.

We used Spearman’s rank correlation^44^ to study the correlation of doubling time and CFR with these indicators. Those that showed consistent trends across all five subsets were then selected for a multivariate regression analysis to analyse the differential impact of each of these factors on Covid19 outcomes in India.

Our analysis showed persistent patterns of socioeconomic vulnerability affecting disease progression - patterns that can provide invaluable support to the designing of evidence-based, locally-responsive interventions and policy.

## 3. Results

Given India’s immense diversity, there is little actionable meaning to “national doubling time” or “national CFR”. For the data to successfully translate into effective intervention, analysis must unearth the localised patterns submerged under national, and even state-level, averaged values. To illustrate this “hidden” variability, we have shown the results for the state of Maharashtra in Figure 2. The red vertical line represents the state average for Maharashtra, while each bar represents the districts. Note the diversity of distribution both within the state and within each district. Similarly, the spectrum of district-level distribution for the entire nation is shown in Figure 3: doubling time ranges from 2–60 days, and CFR from 1% – 28%. The histograms for distributions of average doubling times and average CFR for all districts in India is shown in Suppl. Figure S3. It is therefore vital to perform finer-grained spatial analysis to identify true spread-patterns, and areas that need urgent intervention.

**Figure 2:**
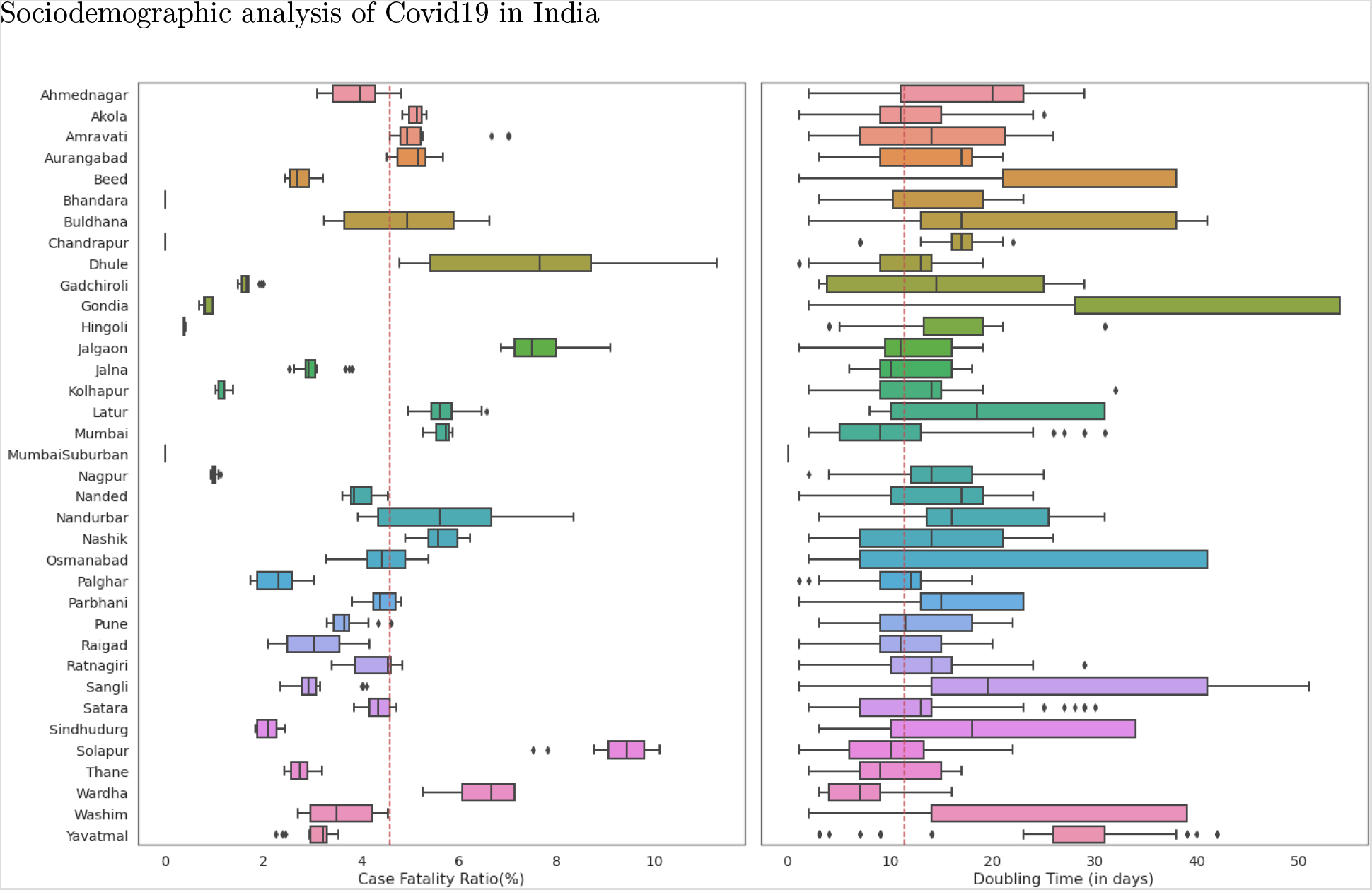
Box plots of variability in Case Fatality Ratio and doubling times for all districts of the state of Maharashtra.

**Figure 3:**
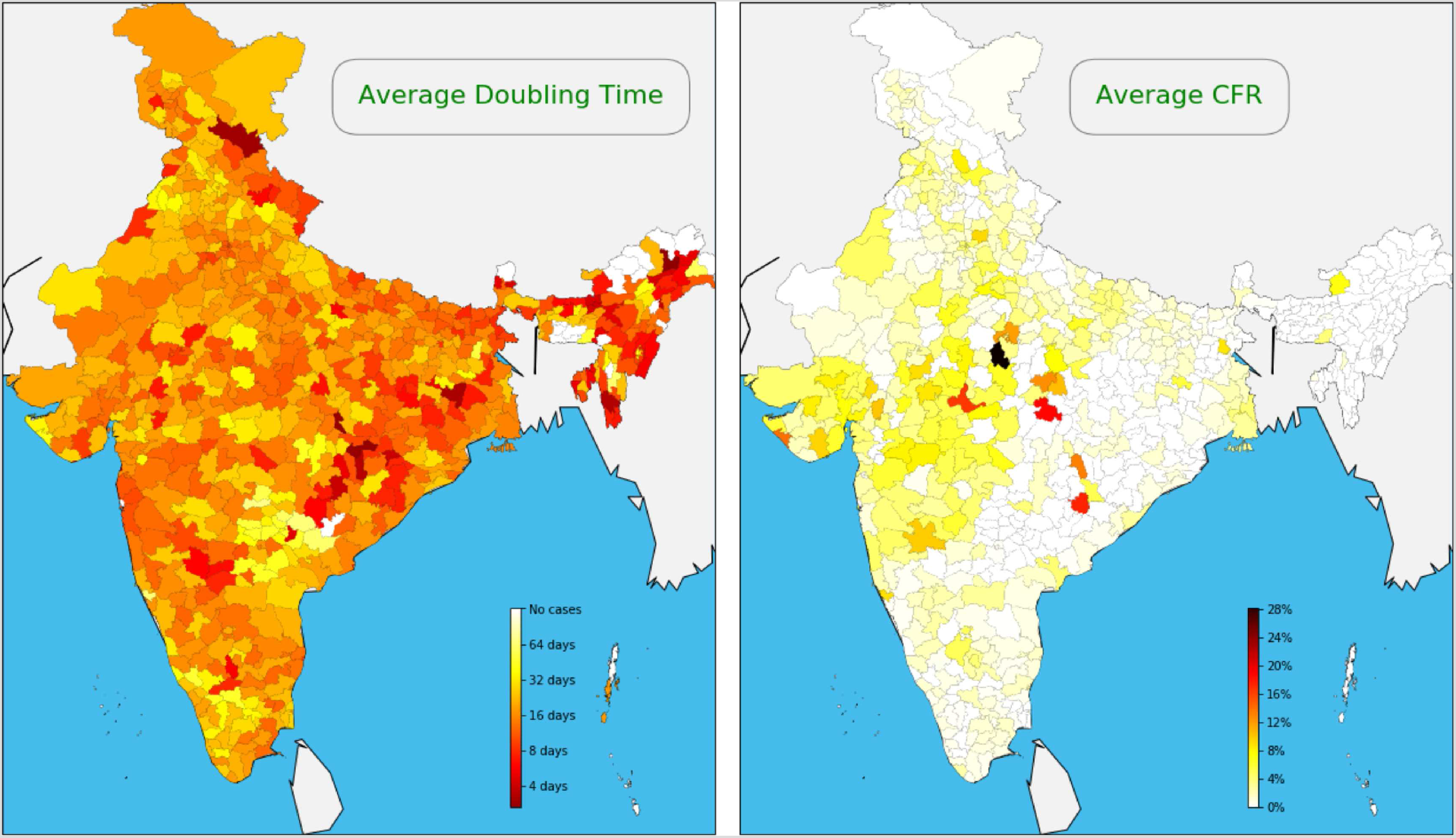
Variability in average doubling times and average CFR at a district level for the entire country. The data is till the 2nd of July, 2020.

We begin with the results of the correlation analysis. For doubling time, we discuss only those eleven correlations where significance p≤0.05 and is consistent across all five subsets. Five sociodemographic indicators are positively correlated with the doubling time, and hence with a slower spread of the disease: (i) literacy rate (ii) proportion of elderly population (iii) population of main workers (iv) proportion of houses in a “good” condition (v) the availability of medical facilities in the district. Conversely, six indicators are negatively correlated with doubling time, and hence with a faster spread of the disease: (i) proportion of children in the district (ii) population of marginal workers (iii) proportion of houses with no drainage (iv) proportion of houses with no latrine (v) proportion of houses with no electricity and (vi) proportion of houses where the source of drinking water was “away”.

For CFR, in order to account for smaller sample sizes, we discuss only those six correlations with a significance p≤0.10 in at least one subset and consistent trends across all five subsets. Of these, three sociodemographic indicators are correlated with low mortality: (i) the availability of medical facilities in the district (ii) the proportion of college graduates (iii) the population of marginal workers. Conversely, these three are correlated with higher CFR: (i) proportion of houses with no latrine, (ii) proportion of houses with no electricity, and (iii) the population of main workers in the district.

A summary of the correlation coefficients of these significant variables, both for doubling time and CFR is shown in Figure 4. The full list of correlation coefficients and significance values are in the Suppl. Table S1 (doubling time) and Suppl. Table S2 (CFR).

**Figure 4:**
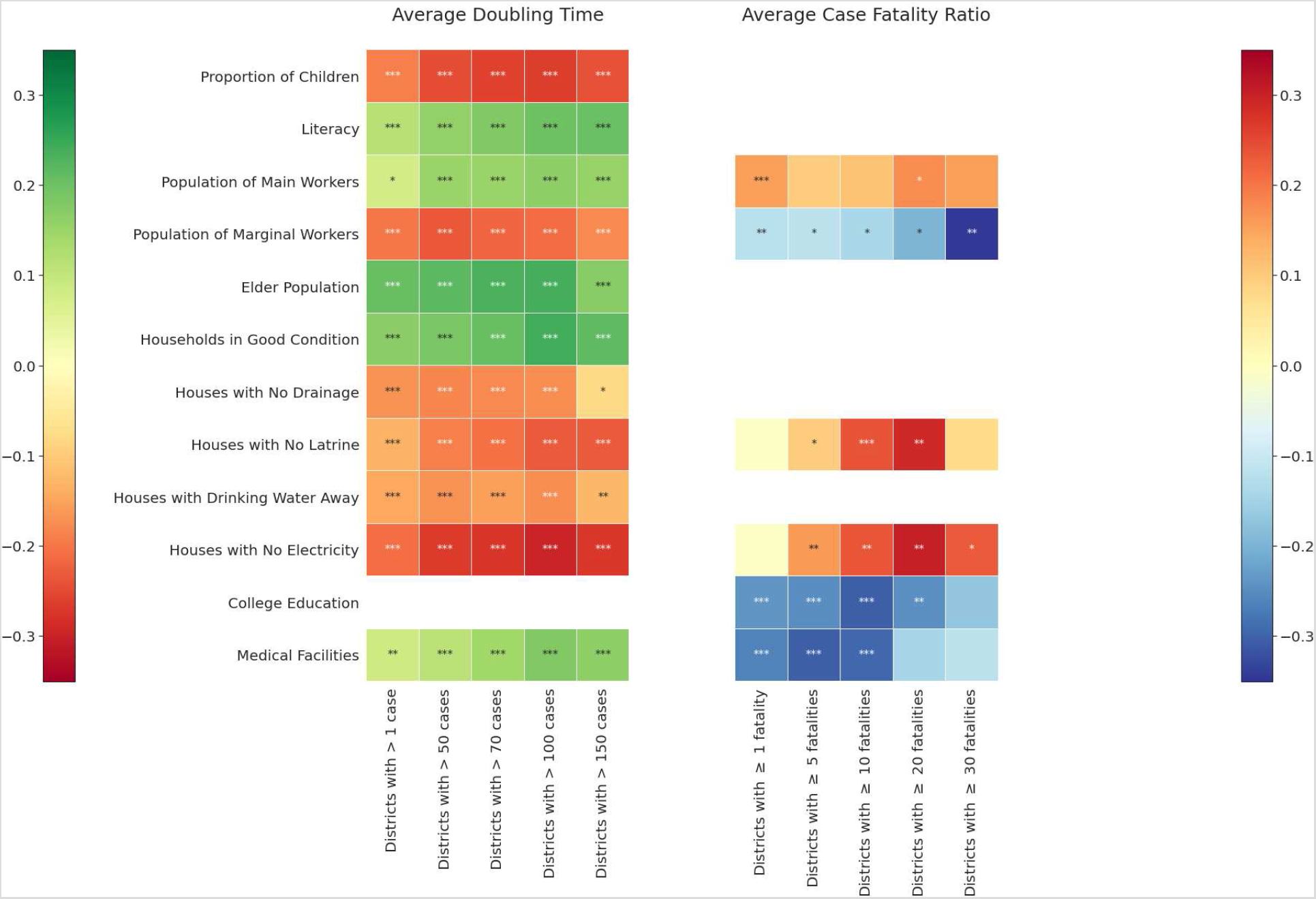
Spearman’s rank correlation coefficient with the average doubling time and CFR. Only those indicators which have consistent correlations across the different subsets have been shown. A single * indicates significance at p ≤ 0.10 (0.20 for CFR), ** indicates significance atp ≤ 0.05(0.10 for CFR), and *** indicates significance at p ≤ 0.01.

We now turn to the results of the multivariate regression analysis. A cross-correlation analysis of the four household indicators indicate that these variables are interdependent and have a high degree of mutual correlation (see Suppl. Figure S4). Therefore, in order to carry out a regression analysis of the doubling time, we chose to retain two of them: (i) “away” source of drinking water (ii) no electricity. Since τ_D_ is a discrete dataset, we use the log of τ_D_ to perform an ordinary linear regression. We also calculated the doubling rate (inverse of the doubling time), and performed a multivariate beta regression^45^ with these nine variables as the rate lies in the (0,1) range. Both analyses yielded identical results. Average doubling time is positively related to the proportion of elder population (significant at p≤0.05), and negatively with number of houses with no electricity (significant at p≤0.01) and a distant source of drinking water (significant at p≤0.05). These trends were consistent across the different subsets analysed.

Since CFR lies in the (0, 1)range, we have constructed a beta regression model with six predictor variables that emerged from the correlation analysis. Two such features- (i) the presence of medical facilities (significant at p≤0.05) and (ii) the proportion of college graduates (significant atp≤0.01), were found to produce low fatality rates. These trends were consistent across the different subsets analysed. Interestingly, in those districts with higher fatalities (> 10 and > 20), we note an emergent pattern of households without electricity or latrines which acts positively to enhance mortality rates (significant at p≤0.05). The results of the beta regression models are summarised in Table 1.

**Table 1:** Coefficients from the beta regression models for the inverse doubling time and CFR. Only those predictors with consistent correlations have been shown. A single * indicates a significant at p ≤ 0.10, ** indicates significant at p ≤ 0.05 and

|  |  | Districts with |  |  |  |
| --- | --- | --- | --- | --- | --- |
| | | $> 1 \text{ case}$ | $> 50 \text{ cases}$ | $> 100 \text{ cases}$ | $> 150 \text{ cases}$ |
| Doubling time | Elder population | -0.14 *** | -0.11 *** | -0.12 ** | -0.06 ** |
|  | Distant drinking water | — | +0.05 * | +0.07 ** | +0.07 ** |
|  | No electricity | +0.11 *** | +0.13 *** | +0.13 *** | +0.11 *** |
|  |  | Districts with |  |  |  |
| | | $\geq 1 \text{ fatality}$ | $\geq 5 \text{ fatalities}$ | $\geq 10 \text{ fatalities}$ | $\geq 20 \text{ fatalities}$ |
| CFR | Medical facilities | -0.24 *** | -0.29 ** | -0.22 * | — |
|  | College graduates | -0.23 *** | -0.24 *** | -0.24 *** | -0.22 ** |

## 4. Discussion

Despite its lower fatality relative to other recent viral epidemics, its high infectivity - coupled with asymptomatic presentation and immunity to current vaccines - has made the containment of SARS-Cov-2 a significant public health challenge. Indeed, prevention seems to rest almost exclusively on maintaining strict physical self-isolation. This is a difficult proposition in general, but particularly so in areas of high population density and poverty^46,47^. India ranks highly on both: its average population density is 382 persons per square kilometre^43^ and nearly all of its population lives on or below $4 a day^48^; globally, it houses 28% of the world’s poor^49^.

The nature of social inequalities, however, is multidimensional. We have therefore chosen to correlate the outcome of Covid19 in India individually to each variable of vulnerability, instead of a composite indicator such as “poverty”.

Five such variables exhibit a persistent pattern of influence on Covid19 outcomes: houses without electricity and/or a proximate source of drinking water correlates with faster spread; a larger elder population correlates with slower spread; a large college-educated population and the presence of medical facilities both correlate with lower mortality.

Cross-correlation analysis of the household variables indicates that absence of electricity and a proximate source of drinking water also relate positively with two other variables: the lack of drainage and of in-house latrines. Collectively, these household-based inequality indicators point to a pattern of multiple daily commutes to, and queuing for, the use of common facilities that are severely cramped, and provide no opportunity for social distancing or disinfection between uses. These four also vary directly with the proportion of marginal workers, further indicating their positive relationship with poverty and the low probability that such households can survive lockdown-unemployment on savings. It should be mentioned here that a significant share of households in India are either one-room (37.1%) or without any exclusive rooms (3.9%)^43^. Between March and June, when Covid19 began its first wave in India in urban centres, social distancing meant that the cross-section of people living in non-electrified houses with a single room or no exclusive room had to stay indoors in darkness and heat - an impossible and inhumane expectation.

While indicators of poverty correlate directly with marginal workers, we should not conflate main workers with stable working or living conditions. A large percentage of main workers are also located on the disadvantaged end of the socioeconomic gradient, with less than $4 a day to survive on^48^. A significant body of recent biosocial research has demonstrated that immersion in unstable environments of scarcity causes a high allostatic load^50,51^, resulting in greater vulnerability to poorer health outcomes^52,53^.

The positive correlation of an elderly population to slower spread is an interesting outcome. While multiple studies have shown that at an individual level, elderly population are at a higher risk of adverse Covid19 outcomes^54^, our analysis suggests that at a population level, a higher elderly population correlates to slower doubling time. It is worth noting that while culturally a large percentage of India’s elderly population still live in large multigenerational families, the explosion of internal migration^55^ has created a pattern of households in which the elderly are “left behind” either seasonally or permanently, and are supported via remittance, savings, or local social networks. This keeps them from needing to participate in the mainstream labour-force, and may have contributed to their lower relative vulnerability vis a vis a younger, working population.

The correlation between the availability of medical facilities and lower CFR is self-evident, however the negative correlation between a college-educated population and CFR merits some attention. Our correlation analysis indicates that college education is the inverse of every indicator of inequality and social marginality we have assessed: lack of electricity, proximate water-source, latrines, and drainage; marginal employment; and larger SC/ST population. In other words, a college education is indexical of relatively privileged socioeconomic position^56^: better access to housing and amenities, stabler income and higher probability of paid leave/work from home, capability to locate public health information, greater likelihood of affording medical care. Thus, the greater the proportion of the socially vulnerable in a district, the lower its probability of a large college-educated populace, and the greater its vulnerability to Covid19 mortality.

## 5. Conclusion

Our work presents a simple statistical association analysis between transmission patterns and outcomes of Covid19 in India, and variables of social vulnerability. We note that our analysis is necessarily ecological and not indicative of individual-level risk. There may potentially be other sociodemographic predictors of disease progression and mortality, and our results are open to potential confounding by these factors. Individual risk-factors for Covid19 - such age, gender, and other comorbidities - have been studied extensively in recent literature^30,57,58^. Our aim was to shift the focus on collective social vulnerabilities and the causes that cause them at the most fine-grained level that the current data would permit us to do, which was at the level of the district. We note that districts in India are themselves large administrative units - and more fine grained analysis can lead to higher correlations with the socioeconomic variables. Our analysis is also hindered by the non-categorisation of Covid19 patient data into existing groups of vulnerability - such as income, caste, gender, rural-urban location, employment status, and so on. With more extensive and complete datasets, it would be interesting to repeat this analysis, perhaps at a ward- or block-level, and see if the qualitative conclusions from the current analysis still hold.

On the subject of data: we have selected our social indicators from the Census of India 2011, which is now a decade old. The profiles of the districts may have undergone changes in the intervening period, potentially affecting results. The disease data has considerable heterogeneity in metrics. Indian states have had differential testing policies, different capabilities for contact tracing and tracking, different definitions Sociodemographic analysis of Covid19 in India for a “Covid19 death”, and different data disclosure policies. Indeed, recent large-scale sero-prevalence studies^59–61^ in Indian cities indicate that Covid19 cases may have been severely underreported, and questions have been raised about the restrictions on enumerating mortality^62,63^.

Our analysis is not without caveats. For assessing transmission, we have used an average of the doubling time of the district. While this provides a model independent characterisation of the transmission dynamics, it must be used with caution. When the pandemic has run its course in a district, the doubling time will grow continuously, and hence including this period in the averaging timespan can inflate the characteristic doubling time. Our data, capped at July 2, 2020, has the advantage of not having seen this period in any district. On the other hand, in the case of CFR, for some districts - especially those with low mortality counts, the average CFR may not be yet reflective of the final value, since the disease is still in its early stages in these districts. We recommend this analysis be redone at the end of the pandemic, when a more comprehensive data is available from every district, to see validated patterns.

Our study validates the hypothesis that inequities embedded in the social structure of the nation affects the dynamics of disease transmission and mortality outcomes. This is particularly relevant in the case of Covid19, because many of India’s early quarantine facilities had unwittingly replicated the very conditions^64,65^ that have been positively correlated with a faster spread (overcrowding, single common source of drinking water, inadequate number of common latrines, common pool of untested and symptomatic populations). Our hope is that this study shall provide policymakers with an evidence-based tool to identify risk-factors specific to a locality and design targeted interventions for them, while also ensuring they cause minimal unintended harm.

More generally, this mode of analysis can be replicated for other infectious diseases in India - chief amongst them tuberculosis and acute diarrhoeic diseases. Diseases, as Rudolf Vichrow pointed out in 1848, is as much a social and political phenomenon as it is clinical^66^, and unearthing the indicators of social vulnerabilities that lie upstream the clinical ones is the first step towards ensuring greater efficiency and public good in the public health system.

## Data Availability

All data is available in the public domain.

## Acknowledgements

The authors acknowledge Prof. Subhankar Karmakar, IIT Bombay for providing the census data for the socio-demographic variables. Helpful discussions within the IIT Bombay Covid19 modeling group are also gratefully acknowledged. MKM acknowledges helpful feedback from Dibyendu Das, IIT Bombay.

## Supplementary materials

**Supplementary Fig.S1.**
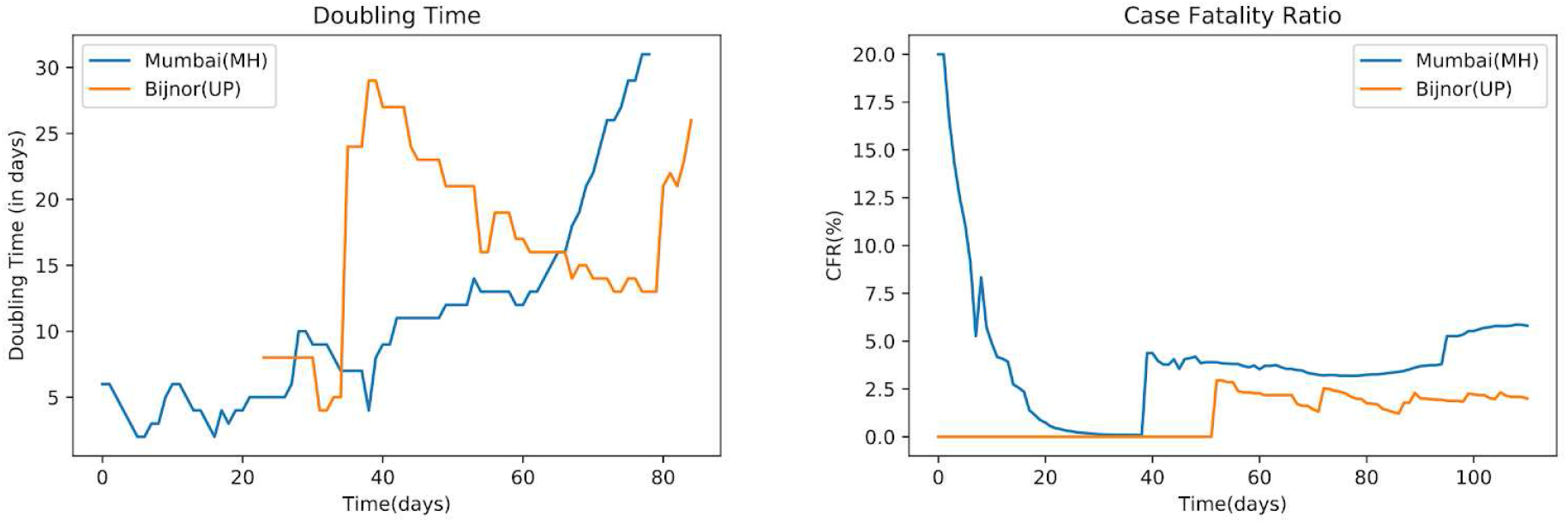
Representative time series plots of the doubling time and the CFR for two districts in India. The two districts shown are Mumbai, Maharashtra and Bijnor, Uttar Pradesh, which have had very different numbers of Covidl9 cases and associated fatalities. Mumbai had 80699 cases and 4689 fatalities, while Bijnor had 300 cases and 6 fatalities, at the time of the analysis. Day 0 represents March 14, 2020.

**Supplementary Fig.S2.**
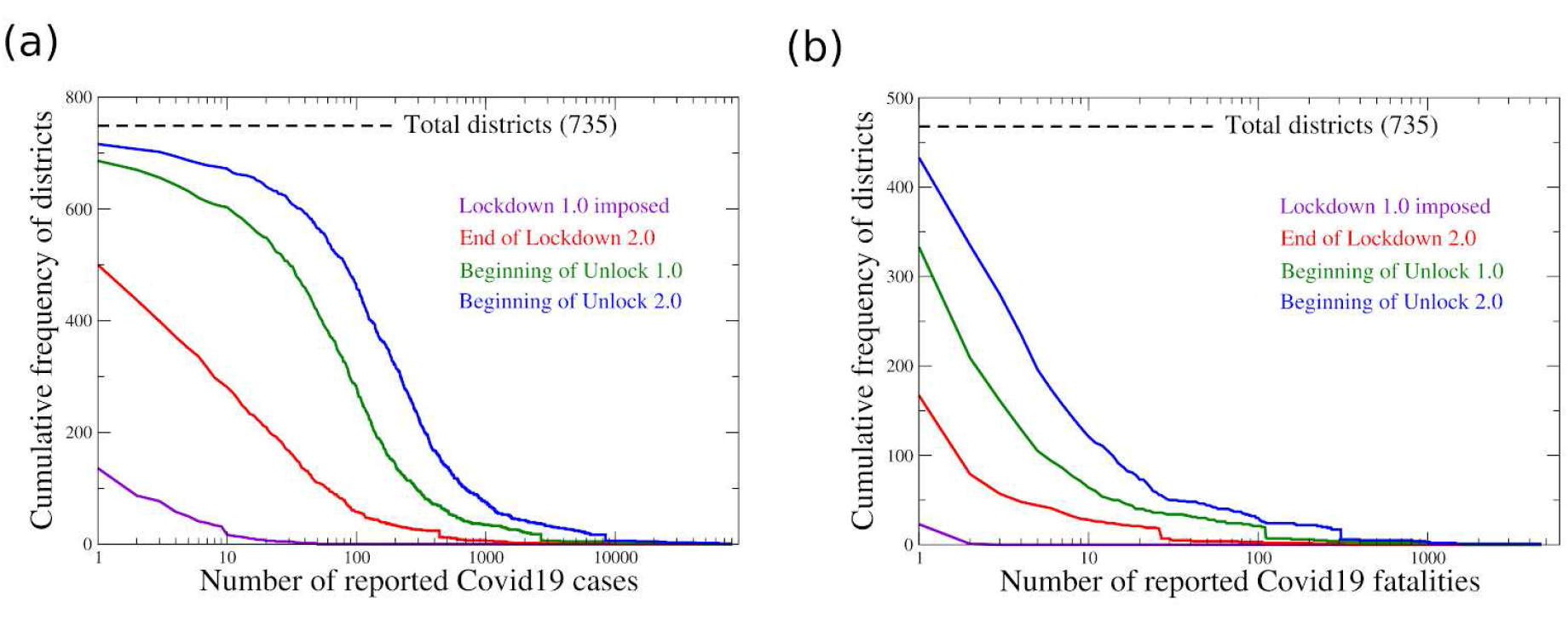
Time evolution of the cumulative frequency distribution of Covidl9 (a) cases and (b) fatalities, across four dates denoting landmarks in India's national containment strategy. Lockdown 1.0 was imposed on March 26th, 2020; Lockdown 2.0 ended on May 3rd, 2020; Unlock 1.0 began on 8th June, 2020; and Unlock 2.0 began on 1st July, 2020.

**Supplementary Fig.S3.**
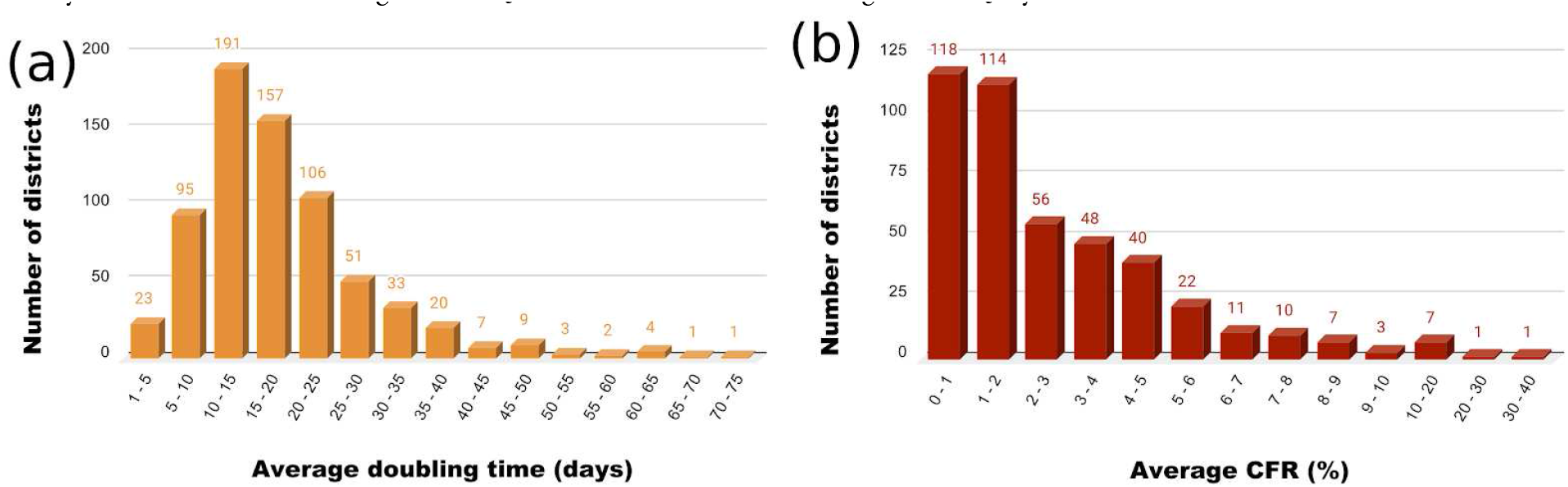
Histograms of (a) average doubling time, and (b) average CFR, for all districts in India.

**Supplementary Table S1:** Full list of Spearman's rank correlation and the corresponding significance values with the doubling time. Correlations with p < 0.05 were considered significant, and are shown in bold (blue).

| Parameters | $\rho_{all}$ | $p_{all}$ | $\rho_{50}$ | $p_{50}$ | $\rho_{70}$ | $p_{70}$ | $\rho_{100}$ | $p_{100}$ | $\rho_{150}$ | $p_{150}$ |
| --- | --- | --- | --- | --- | --- | --- | --- | --- | --- | --- |
| Total Households | **0.13** | <0.01 | 0.08 | 0.08 | 0.05 | 0.31 | -0.00002 | 0.99 | 0.04 | 0.44 |
| Total Population | **0.11** | 0.01 | 0.05 | 0.27 | 0.02 | 0.73 | -0.03 | 0.49 | 0.003 | 0.95 |
| Children | **-0.19** | <0.01 | **-0.24** | <0.01 | **-0.26** | <0.01 | **-0.26** | <0.01 | **-0.24** | <0.01 |
| SC-ST | **-0.09** | 0.02 | -0.04 | 0.34 | -0.03 | 0.47 | -0.03 | 0.59 | -0.05 | 0.33 |
| Literacy | **0.12** | <0.01 | **0.16** | <0.01 | **0.18** | <0.01 | **0.20** | <0.01 | **0.20** | <0.01 |
| Main Workers | **0.08** | 0.06 | **0.15** | <0.01 | **0.15** | <0.01 | **0.16** | <0.01 | **0.16** | <0.01 |
| Marginal Worker | **-0.20** | <0.01 | **-0.23** | <0.01 | **-0.22** | <0.01 | **-0.21** | <0.01 | **-0.18** | <0.01 |
| Non Work | 0.04 | 0.38 | -0.02 | 0.6 | -0.04 | 0.42 | -0.06 | 0.24 | -0.06 | 0.26 |
| Elder Population | **0.20** | <0.01 | **0.22** | <0.01 | **0.23** | <0.01 | **0.24** | <0.01 | **0.17** | <0.01 |
| Disability Population | -0.02 | 0.67 | -0.02 | 0.72 | -0.02 | 0.70 | -0.06 | 0.21 | **-0.12** | 0.03 |
| Good HH Condition | **0.17** | <0.01 | **0.19** | <0.01 | **0.21** | <0.01 | **0.24** | <0.01 | **0.22** | <0.01 |
| Bad HH Condition | -0.004 | 0.92 | -0.05 | 0.31 | -0.07 | 0.16 | **-0.13** | 0.01 | **-0.16** | <0.01 |
| No Drainage | **-0.17** | <0.01 | **-0.18** | <0.01 | **-0.18** | <0.01 | **-0.17** | <0.01 | **-0.08** | 0.16 |
| No Latrine | **-0.13** | <0.01 | **-0.19** | <0.01 | **-0.20** | <0.01 | **-0.23** | <0.01 | **-0.23** | <0.01 |
| Drinking water away | **-0.15** | <0.01 | **-0.17** | <0.01 | **-0.16** | <0.01 | **-0.17** | <0.01 | **-0.13** | 0.02 |
| No Electricity | **-0.21** | <0.01 | **-0.27** | <0.01 | **-0.28** | <0.01 | **-0.30** | <0.01 | **-0.27** | <0.01 |
| Primary Education | **-0.12** | <0.01 | **-0.10** | 0.03 | -0.08 | 0.11 | -0.04 | 0.40 | -0.05 | 0.42 |
| Secondary Education | 0.01 | 0.81 | 0.06 | 0.20 | 0.08 | 0.10 | **0.10** | 0.05 | 0.03 | 0.60 |
| College | -0.02 | 0.61 | 0.02 | 0.64 | 0.02 | 0.64 | 0.05 | 0.32 | 0.06 | 0.25 |
| Medical Facility | **0.09** | 0.04 | **0.11** | 0.01 | **0.14** | <0.01 | **0.18** | <0.01 | **0.16** | <0.01 |

**Supplementary Table S2:**
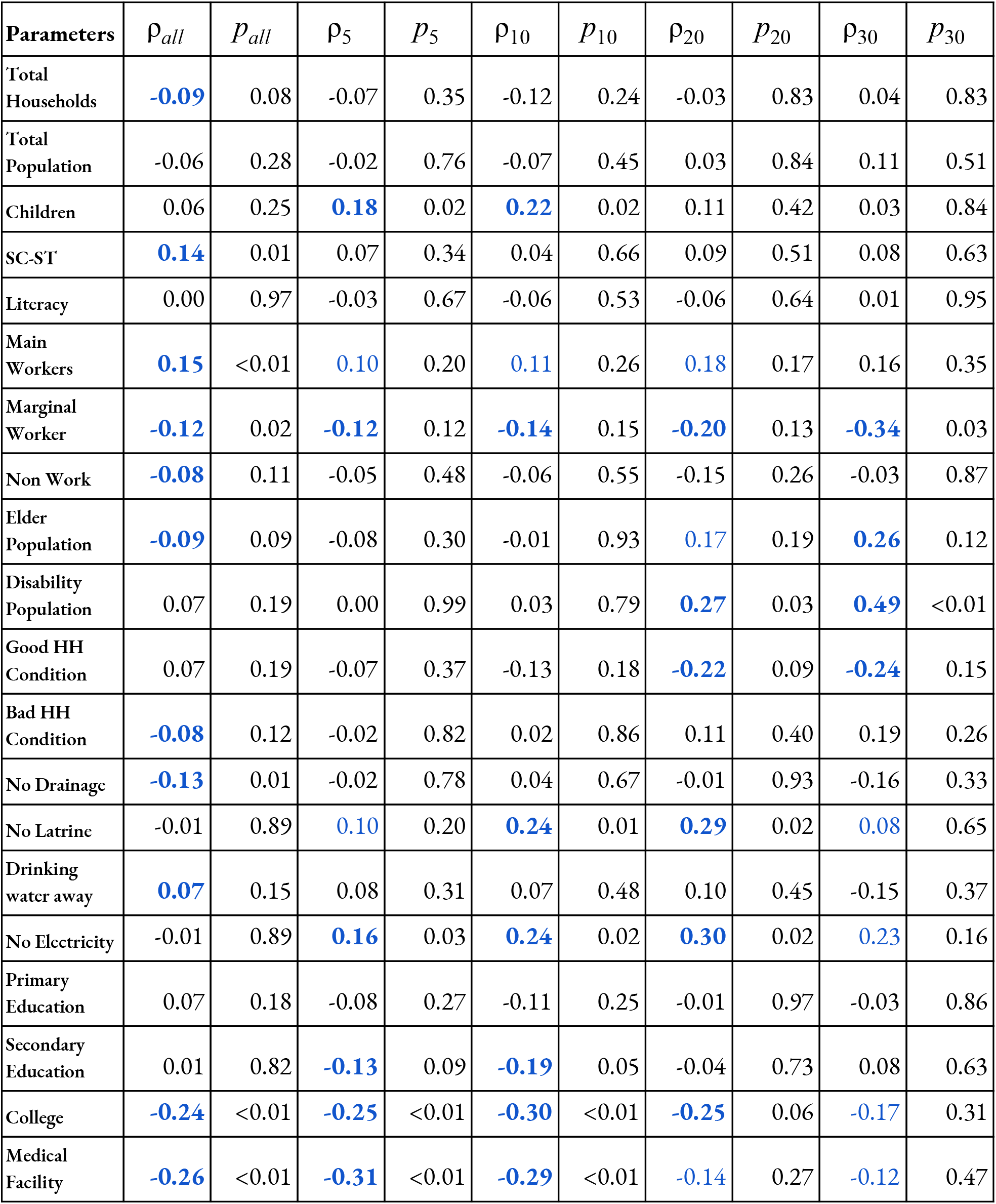
Full list or Spearman s rank correlation and the corresponding significance values with the CFR. Correlations with p < 0.15 were considered significant, and are shown in bold (blue).

**Supplementary Fig.S4.**
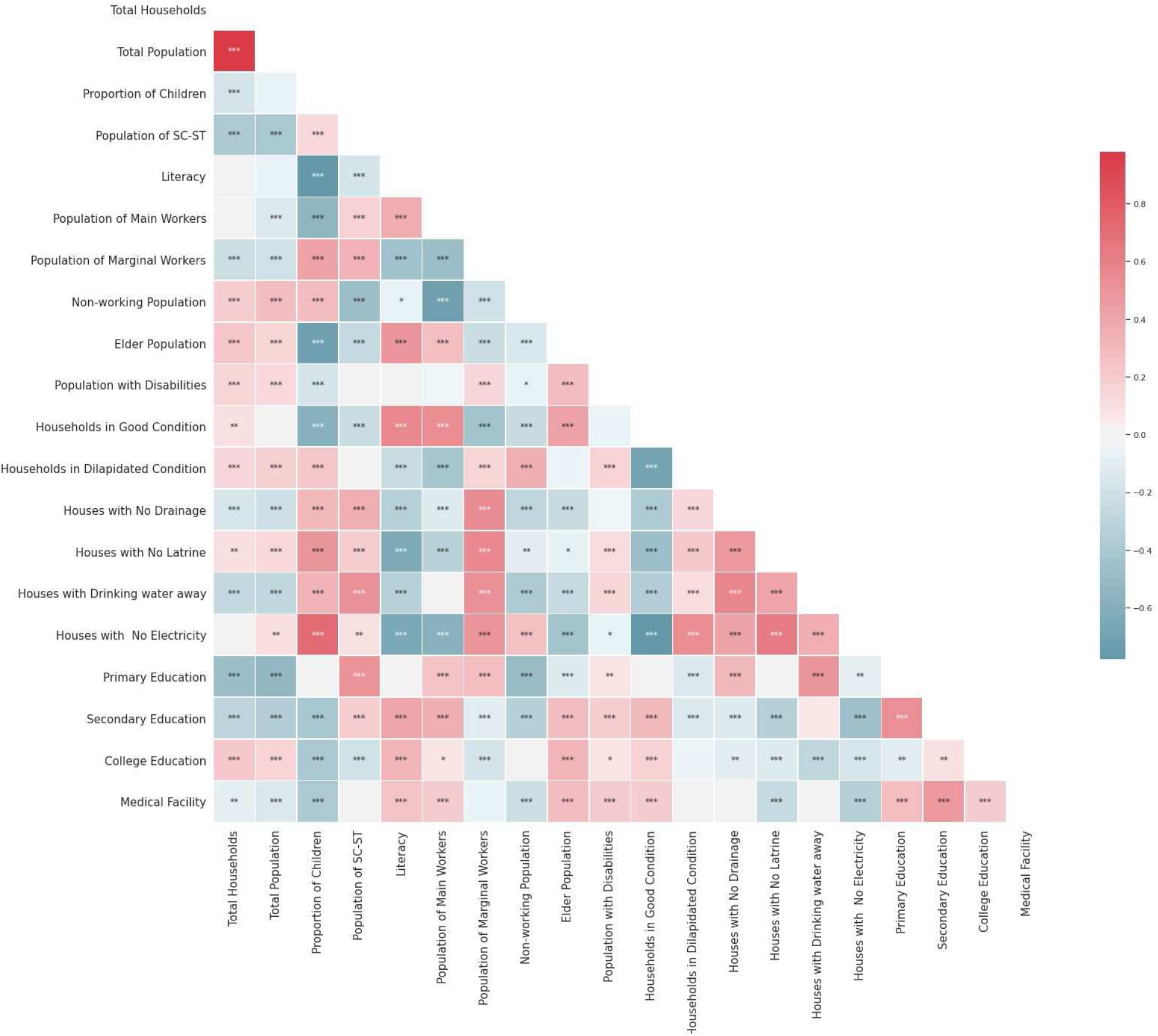
Cross-correlations of the twenty chosen vulnerability indicators. The significance of the corresponding Spearman's correlation coefficients are printed inside the respective square.

